# FHIR “Profiliferation”: A Data Science Approach

**DOI:** 10.1101/2022.03.09.22272163

**Authors:** Mark A. Kramer

## Abstract

Profiling is a mechanism for shaping Fast Healthcare Interoperability Resources (FHIR) for particular use cases. “Profiliferation” (profile + proliferation) is a coinage referring to the explosive growth in the number of FHIR profiles over the past few years. By reviewing a broad sample of almost 3000 FHIR profiles from 125 implementation guides, it was determined that just two items, Observation and Extension, accounted for half the profiles. FHIR’s 80/20 rule was determined to be closer to 65/35, revealing that FHIR is more dependent on profiling than initially intended. Use of the Observation resource was inconsistent, hinting that the resource is either poorly designed, or that users lack proper guidance. Better management of reusable items and certain changes in FHIR and profiling practices could improve the consistency of FHIR artifacts and reduce unnecessary and potentially incompatible profiles.

## Introduction

Health Level 7 Fast Healthcare Interoperability Resources ^1^ (HL7® FHIR®) is a framework for interoperability. FHIR defines a data exchange syntax, an application programming interface (API), a set of health-related resources (information objects), some terminology, and an extensibility mechanism. By design, FHIR omits many details needed to implement health-related use cases.

Those details are provided by Implementation Guides (IGs) ^2^. Among the most important items in IGs are FHIR profiles ^3^. Profiles provide additional information for the use case’s data structures, including constraints on data element cardinality, value sets, additional data elements (“extensions”), and more. Profiles can also be used at run time to check the validity of exchanged data.

Over the past few years, the FHIR community has built thousands of profiles ^4^, leading to challenges in discovery and re-use of these resources. Given the difficulties of finding existing profiles, scarce knowledge of their provenance, and lack of control over third party development, IG authors frequently elect to create their own profiles rather than reusing existing ones, thus compounding the problem.

Before formulating approaches to deal with profile proliferation (“profiliferation”), it is worthwhile to analyze what specifically is being done with profiles. By studying extant profiles, it is possible to transform the problem from simply “too many profiles” to one that spotlights specific areas and practices that can be addressed.

## Methodology

The current study is a meta-analysis of the contents of IGs to identify patterns of profile specification. The sample of IGs was selected from the population of IGs listed in the FHIR Package Registry ^5^. When accessed in February 2022, the Registry contained over 450 distinct IG packages based on FHIR Release 4 (R4). Eliminating packages uploaded for testing purposes, those created prior to 2020, and those containing serious errors, 125 IGs were selected (Appendix A). The majority of these IGs are cross-listed in the FHIR Implementation Guide Registry ^6^, a more selective source that indicates a degree of maturity in the content of those IGs.

The selected IGs were broad-based. A total of 11 countries were represented, the most numerous being 53 IGs from the US, 24 from Germany, 9 from Switzerland, and 5 from Canada, with 24 considered universal (not country-specific). The domains covered by the IGs included financial management, public health, administration, patient care, quality measurement, clinical decision support, clinical documents, medical records access, medications, and more.

The StructureDefinitions ^7^ in each IG were analyzed. StructureDefinition is the FHIR resource used to define profiles of resources and datatypes. A small number of StructureDefinitions defining logical models (custom resources) were excluded. Other FHIR conformance resources ^8^such as ValueSet, OperationDefinition, and SearchParameter were left out of scope.

StructureDefinitions contain a “differential” that captures all the changes made by the profile, relative to the profile’s immediate parent. These changes include adding new data elements, restricting cardinality (for example changing an optional element to a required element), assigning value sets to data elements, and constraining datatypes.

For each StructureDefinition, a “deep differential” was constructed by recursively combining the differential of the target profile with the differentials of its parents (Figure 1). The deep differential captures every modification made by the target profile on its base resource or datatype. Finally, for each data element, the types of profiling modifications were noted.

**Figure 1.**
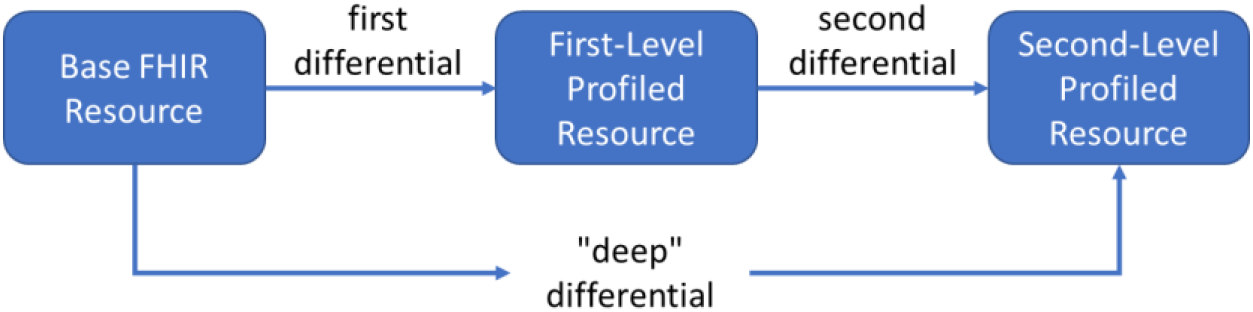
Formulation of the deep differential across multiple levels of profiles (generalizes to any depth).

### Classification of Data Elements

To help organize and interpret the results, the data elements were subdivided into three categories:

1. **Native Elements**. These are elements defined in the core FHIR specification. Native elements include an element’s sub-elements to any depth (whether explicit or implicit) and any inherited data elements. For example, the element Patient.address implies the nested native elements Patient.address.city and Patient.address.postalCode. Inheritance accounts for implicit elements such as Patient.meta.language. Native elements meant to contain extensions, such as Patient.extension, and elements of the Extension datatype, such as Extension.value[x], are also considered native. *Top-level elements* are those native elements that have no parent element but the resource itself.
2. **Slices and Slice Elements**. Slicing is used to introduce constraints that do not necessarily apply to the whole element, most often an array (repeated) element. For example, a profile author might define a blood pressure profile by slicing the Observation.component array element into two slices: Observation.component:SystolicBP and Observation.component:DiastolicBP, each slice following different constraints. *Slice elements* include the sliced element and of its sub-elements, for example, Observation.component:SystolicBP.valueQuantity. The number of slice elements is naturally greater than the number of slices.
3. **Extensions and Extension Elements**. FHIR allows user-defined extensions on resources and datatypes ^9^. Extensions are used to define new data elements that are missing from the base specification. For example, the Canadian baseline IG adds a new element for classifying the person’s aboriginal identity, Patient.extension:aboriginalidentitygroup. *Extension elements* include the extension and all its sub-elements, for example, Patient.extension:aboriginalidentitygroup.valueCoding.system. Technically, extensions are slices on extension elements, but for the purposes of this study, they are different enough to be considered separately.

There are two types of extensions: standalone and inline ^10^. Standalone extensions are defined by profiles of the Extension datatype, while inline extensions are defined without reference to a separate Extension profile. Inline extensions are used in complex extensions (extensions of extensions).

It is possible for a single data element to involve both slicing and extension. For example, the element Patient.identifier:JHN.extension:versionCode, combines a slice on Patient.identifier representing the Jurisdictional Health Number (JHN) with an extension representing the version of the JHN. For the purposes of this study, elements involving both slices and extensions were classified as extensions.

Regarding the subdivision of elements into these three categories, there might be an argument to classify slices as native since they are constraints on native elements. However, the identical argument would also apply to extensions, which are constraints on native extension elements. It is more informative to view slices as their own category, falling between the more rigid, predefined meaning of some native elements and the flexible semantics of extensions. For example, Observation.component is composed of arbitrary name-value pairs, like extensions, but the semantics are more constrained, since each component is supposed to represent an observation concurrent with the primary observation.

It is worth noting that native elements are much more numerous than the FHIR documentation indicates. For example, the documentation page for the Patient resource lists 28 data elements ^11^, while the StructureDefinition contains 73 ^12^, yet this study observed 126 distinct native data elements. The difference is that neither the documentation nor the StructureDefinition fully accounts for nested and inherited elements.

## Results

### Number of Profiles, Slices, and Extensions

The 125 selected IGs contained a total of 2,931 profiles. The IG with the most profiles (hl7.fhir.us.covid19library) had 150, while the median was 16 profiles, and the average 23.5 profiles. About one-third of the IGs (43 of 125) had 10 or fewer profiles.

The 15 most frequently profiled resources and datatypes are shown in Table 1. Overall, 105 distinct FHIR resources and 13 complex datatypes were profiled. The remaining 40 resources and 50 datatypes defined in FHIR R4 were never profiled. Of the 40 unprofiled resources, 27 were “draft” status (FHIR Maturity Model ^13^ level 0), indicating very little use in the FHIR community. Only two unprofiled resources were maturity level 3 or higher (AppointmentResponse and OperationDefinition).

**Table 1.** Most frequently profiled resources and data types, with number of slices and extensions per resource type.

| Rank | Profiled Item (Resource or Datatype) | Number of IGs where the item appears | Total Number of Profiles of Item | Total Number of Slices in All Profiles | Total Number of Extensions in All Profiles |
| --- | --- | --- | --- | --- | --- |
| 1 | Extension | 89 | 936 | 13 | 512 |
| 2 | Observation | 59 | 502 | 1763 | 340 |
| 3 | Bundle | 54 | 131 | 298 | 0 |
| 4 | Composition | 41 | 85 | 1295 | 227 |
| 5 | Condition | 40 | 82 | 82 | 91 |
| 6 | Patient | 61 | 74 | 144 | 302 |
| 7 | Organization | 51 | 67 | 183 | 121 |
| 8 | Practitioner | 46 | 53 | 112 | 54 |
| 9 | Location | 29 | 36 | 12 | 15 |
| 10 | ServiceRequest | 28 | 35 | 32 | 57 |
| 11 | Procedure | 24 | 33 | 28 | 66 |
| 12 | Identifier | 9 | 31 | 0 | 0 |
| 13 | Device | 21 | 26 | 38 | 21 |
| 14 | Task | 13 | 26 | 40 | 36 |
| 15 | Coverage | 21 | 25 | 30 | 20 |
| **Total (all resources/datatypes)** |  | **125** | **2931** | **4864** | **2645** |

In total, there were 4864 slices defined in the profiles, about 1.7 per profile. The ten most frequently sliced elements were: Composition.section (1,164 slices), Observation.component (534 slices), Observation.identifier (382), Observation.performer (373), Bundle.entry (291), Observation.category (212), Organization.identifier (139), Observation.hasMember (88), Observation.code.coding (87), and Patient.identifier (84).

In total, the IGs used extensions 2645 times (Figure 2). Of these, 2,291 were standalone extensions and 354 were inline extensions. Of the standalone extensions, 526 (23%) were standard extensions (included in base FHIR).

**Figure 2.**
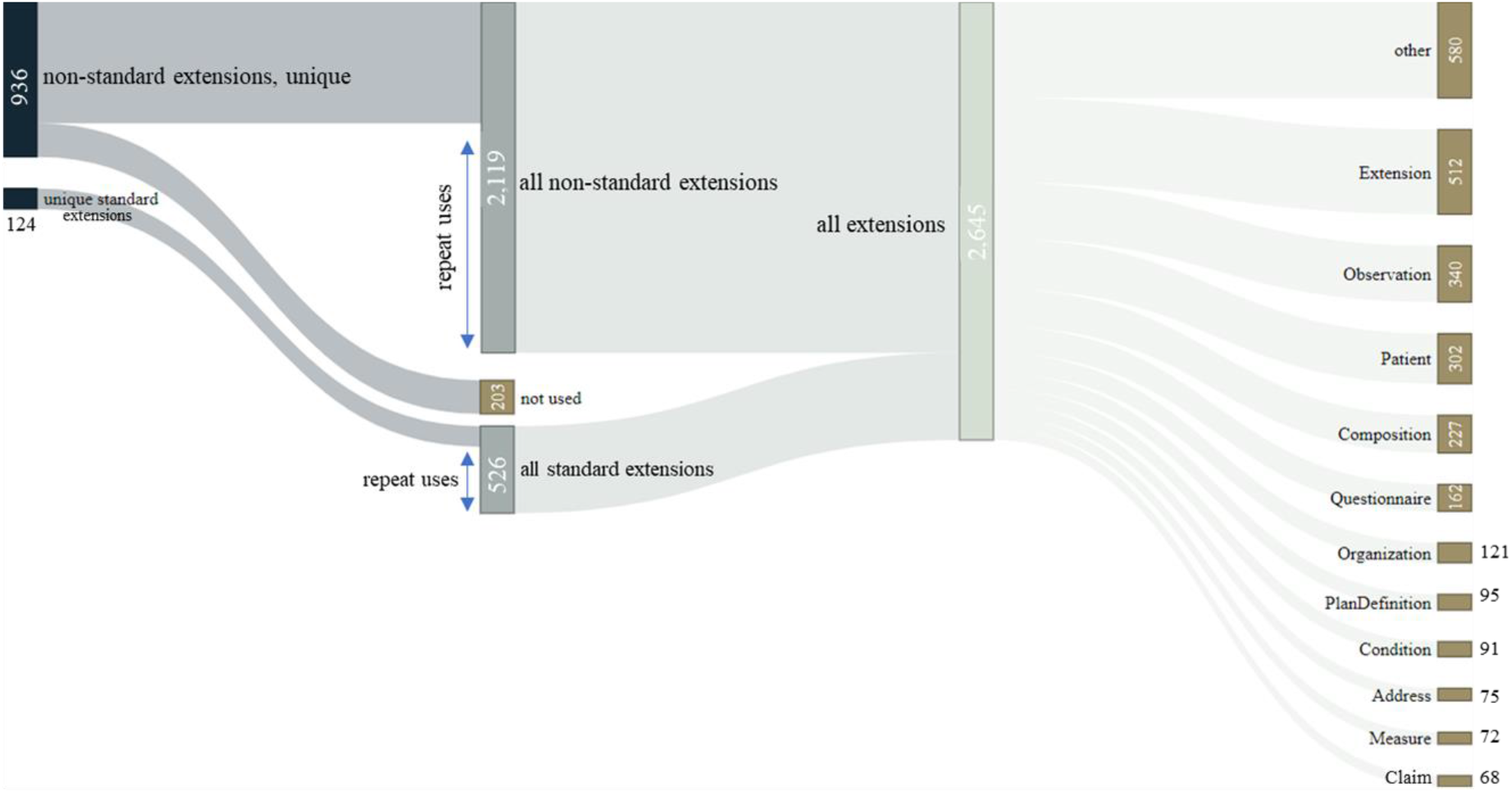
Sources and usage of extensions in the sample IGs.

The standard extensions represented 124 unique extensions. The ten most frequently used standard extensions were: data-absent-reason (used 49 times), condition-assertedDate (42), bodySite (24), workflow-episodeOfCare (18), designNote (18), observation-secondaryFinding (14), narrativeLink (13), patient-birthPlace (12), rendering-style (11), and rendering-xhtml (11).

The ten most frequently extended resources were: Extension (512 times), Observation (340), Patient (302), Composition (227), Questionnaire (162), Organization (121), PlanDefinition (95), Condition (91), Address (75), Measure (72), and Claim (68). Note that extending Extension is how complex (nested) extensions are created.

### Profiling Actions

Multiple profiling actions can be applied to one data element. Ten of the most common profiling actions are: changing the minimum or maximum cardinality, assigning a “must support” (MS) flag, constraining the datatype, assigning a fixed or pattern value, binding a value set or changing the binding strength, and adding slice(s) or extension(s). One of more of these actions were taken on each of the 52,397 profiled data elements across all IGs. Other profiling actions, such as adding descriptions, providing metadata about the profile, specifying upper and lower bounds on quantitative elements, etc., were not tracked. Table 2 shows the percentage of profiled data elements profiled each way.

**Table 2.** Frequency of ten common profiling actions across profiled native elements, extensions, and slices.

|  | All Profiled Data Elements | Profiled Native Elements | Profiled Slice Elements | Profiled Extension Elements |
| --- | --- | --- | --- | --- |
| Tag as "MustSupport" | 41% | 31% | 61% | 53% |
| Change Minimum Cardinality | 39% | 41% | 37% | 36% |
| Change Maximum Cardinality | 38% | 32% | 45% | 53% |
| Constrain Datatype | 27% | 25% | 21% | 55% |
| Change the Binding Strength | 8% | 6% | 11% | 11% |
| Assign a Value Set | 8% | 9% | 5% | 4% |
| Add Slice(s) | 7% | 9% | 5% | 4% |
| Require a fixed value exactly | 6% | 2% | 15% | 1% |
| Add Extension(s) | 6% | 8% | 4% | 3% |
| Require a value pattern | 4% | 4% | 5% | 3% |

### Must Support Elements

Labeling an element “must support” is significant because it indicates the data element is of particular importance in the IG ^14^. Assigning these flags is the most common profiling action. Table 3 shows which native elements were most likely to be MS, for the top 15 resources and data types. Top level elements not listed in the Table 3 are rarely designated MS in IGs and perhaps could have been omitted from the resource in question. It may be noted, however, that some IGs do not specify MS flags.

**Table 3.**
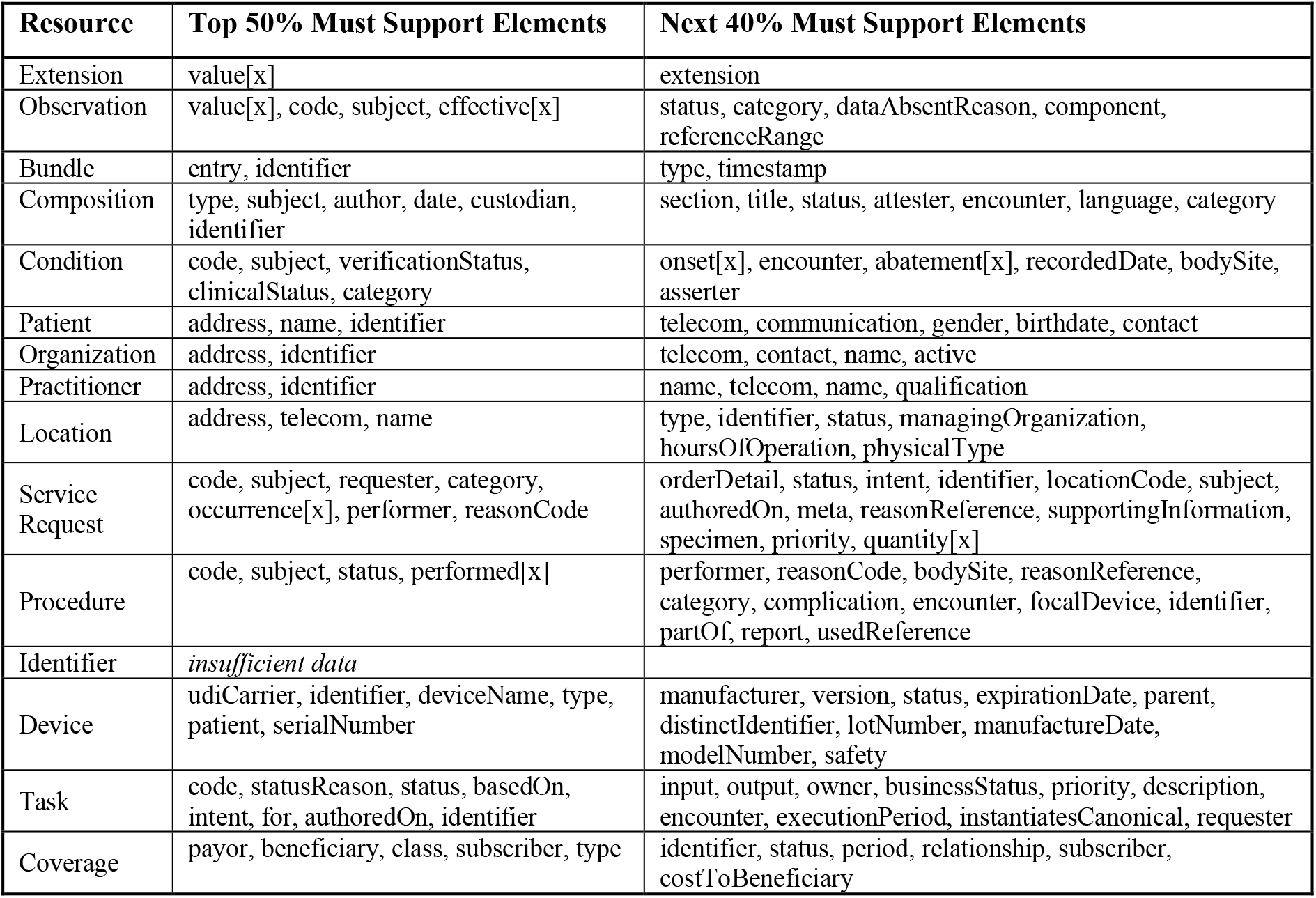
Data elements most frequently flagged as MS, by resource, in order of frequency, rolled up to the top level by summing counts of MS sub-elements. Elements in the bottom 10% are not listed.

### Evaluating FHIR’s 80/20 Rule

The native data elements in a FHIR resource are intended to cover most use cases. FHIR’s 80/20 rule ^15^ states that FHIR covers 80% of data elements in existing healthcare systems, leaving 20% to be dealt with as extensions. There are also caveats that this rule is not intended to be a statistical measure, only a rule of thumb. Nonetheless, it is important to know to what extent base FHIR is capturing the most of the important data elements needed in IGs.

Assuming MS elements are those most important to a use case, the percentage of native MS elements versus all MS elements can provide experimental confirmation of the 80/20 rule. The rule turns out to be true for some resources, but not others (Table 4). Condition, Practitioner, Location, ServiceRequest, and Device exceed 80%, while Observation, Patient, Organization, Procedure, Task, and Coverage fall short, signaling that on average, users have been customizing FHIR more than hoped. Because not all IGs specify MS flags, the last column of Table 4 shows the same calculation based on all elements profiled across all IGs. The results are similar, but several more resources fall below the 80% line. According to this analysis, FHIR’s 80/20 rule is more like 65/35.

**Table 4.** Evaluation of FHIR’s 80/20 rule according to the percentage of native (non-slice, non-extension) MS elements in all MS elements, by resource. For comparison, the percentage based on all profiled elements also shown.

|  | Count of All MS Elements (all IGs) | Count of Native Elements Among All MS Elements | % Native Elements Among All MS Elements | % Native Elements Among All Profiled Elements |
| --- | --- | --- | --- | --- |
| Location | 240 | 215 | 89.6 | 86.4 |
| Practitioner | 442 | 376 | 85.1 | 59.3 |
| Device | 148 | 125 | 84.5 | 65.5 |
| ServiceRequest | 489 | 401 | 82.0 | 88.9 |
| Condition | 739 | 594 | 80.4 | 80.8 |
| Patient | 943 | 706 | 74.9 | 60.5 |
| Organization | 753 | 560 | 74.4 | 58.2 |
| Identifier | 7 | 5 | 71.4 | 96.5 |
| Observation | 3769 | 2686 | 71.3 | 60.0 |
| Procedure | 315 | 220 | 69.8 | 74.1 |
| Task | 463 | 322 | 69.5 | 72.0 |
| Coverage | 210 | 145 | 69.0 | 71.4 |
| Bundle | 631 | 370 | 58.6 | 56.5 |
| Extension | 478 | 138 | 28.9 | 6.2 |
| Composition | 3342 | 594 | 17.8 | 23.9 |
| **Total (all resources)** | **20659** | **13434** | **65.0** | **62.2** |

### Observation Profiles

As the most profiled resource, Observation deserves special attention. Table 5 shows which top-level elements were profiled most frequently, with each count including profiled sub-elements. Observation.component was by far the most frequently profiled element because numerous slices created on this element.

**Table 5.** Profiling of elements in the Observation resource, rolled up to top-level elements. Unlisted elements were profiled less than 100 times.

| Top-Level Data Element | Count of All Profiled Elements | Profiled Native Elements | Profiled Slice Elements | Profiled Extension Elements |
| --- | --- | --- | --- | --- |
| component | 2684 | 531 | 1951 | 202 |
| value[x] | 1052 | 1023 | 24 | 5 |
| code | 982 | 647 | 255 | 80 |
| identifier | 976 | 183 | 793 | 0 |
| category | 702 | 354 | 348 | 0 |
| performer | 685 | 312 | 373 | 0 |
| subject | 569 | 569 | 0 | 0 |
| effective[x] | 416 | 409 | 0 | 7 |
| dataAbsentReason | 341 | 320 | 21 | 0 |
| status | 329 | 320 | 0 | 9 |
| bodySite | 295 | 257 | 12 | 26 |
| method | 248 | 240 | 0 | 8 |
| extension | 234 | 80 | 0 | 154 |
| referenceRange | 192 | 166 | 25 | 1 |
| hasMember | 173 | 85 | 88 | 0 |
| interpretation | 133 | 104 | 25 | 4 |
| meta | 132 | 104 | 28 | 0 |
| device | 104 | 104 | 0 | 0 |
| specimen | 104 | 104 | 0 | 0 |

Focusing on Observation.component, what stands out is that many components defined in profiles do not appear consistent with that element’s definition. Components are supposed to be observations that accompany or group with the primary observation. However, in practice, components were used to capture a wide variety of information, including circumstances surrounding the observation, methods, follow-up actions, interpretations, and more. Without judging the appropriateness of any particular one, here are some examples of observation components (names formatted for readability):

- Administered Activity
- Assay Code Radioisotope
- Assigning Authority
- Birth Year
- Case ID
- City
- Conclusion String
- Coverage Code
- Daily Insulin Dose
- Endoscopically Removed
- Evidence Level
- Measurement Confidence
- Measurements Per Day
- Molecular Consequence
- Pack Years
- Procedure Reported
- Prognosis
- Region Studied
- Reporting Timeframe
- Route of Administration
- Sent to Pathologist
- Travel Start/End Date
- Underlying Medical Condition

The variety of top-level extensions was also conspicuous, presenting things that might align with certain elements in the Observation resource, or expressed in another resource. Here are examples of top-level extensions in Observation:

- Assistance Required
- Associated Precondition
- Assigning Authority
- Body Position
- Body Structure
- Case Feature Pertinence
- Clothed During Measurement
- Current Job
- Death Location
- Disclaimer
- Disposition Location
- Employer
- Episode of Care
- Event Location
- Evidence Type
- Exercise Association
- Exposure Type
- Injury Location
- Lab Test Priority
- Measurement Device
- Not Done Reason
- Precision Value
- Pregnancy Status Date
- Prescription
- Recommended Action
- Reportability
- Sensor Description
- Secondary Finding
- Sleep Status
- Treatment Plan

Some other observations:

- Some Observations with many components appeared to be more like questionnaires, with a series of questions and answers.
- When profiling elements such as category, identifier, code, and performer, users were split between using fixed values, slicing, and value pattern approaches.

## Discussion

Base FHIR provides approximately 2/3 of the necessary elements in the IGs studied here. One reason for profiliferation is that profiling is used to define 33-38% of new data elements, rather than the 20% suggested by FHIR’s 80/20 rule.

With almost 3000 profiles gathered from 125 IGs, only two profiled items, Extension and Observation, accounted for nearly half (49%) of all profiles. These are the prime targets for reducing profiliferation.

Although reusable “standard extensions” are included with FHIR, they accounted for only one-fifth of all extensions used in the IGs. There are clearly opportunities to increase this proportion. Currently, standard extensions are only updated when a new FHIR version is released. Release can be several years apart, which is far too slow for IG authors.

An alternative source of existing extensions is Simplifier.net ^4^, which currently contains about 1500 active extensions for R4. A major drawback of Simplifier is that it is not curated, so its contents come with no guarantee of quality, stability, or uniqueness. There is no standardization when users can choose from 31 birth sex extensions.

Since medicine has many thousands of potential observables, extensive profiling of Observation is to be expected. However, it is apparent from this survey that the use of this resource is inconsistent. This has been the subject of previous discussions ^16^. It could be that the resource is poorly designed, or that users lack proper guidance. Regardless, if the current situation persists, implementers will be faced with adapting their input and output processing to deal with many idiosyncratic Observation profiles.

Although not a direct source of profiliferation, slices are a major contributor to variation within profiles. In addition, slices are even less reusable than extensions because they are typically defined inline. To allow any reuse, the user must first create a standalone profile of the required datatype, and then constrain the slice datatype to an instance of that profile. However, this approach does not apply to BackboneElement ^17^, a hierarchical grouping element where slicing often occurs. FHIR does not allow profiling of nested structures based directly on BackboneElement, such as Observation.component or Patient.contact, and yet one can profile very similar elements, such as Observation.identifier or Patient.address. The difference is that the latter are based on normally-behaving data types (Identifier and Address, respectively). Inability to profile BackboneElement structures limits the reusability of slices.^1^

## Recommendations

Based on the current study, the following recommendations may help reduce profiliferation:

1. Evaluate what data elements should be added to each resource, so the coverage of native elements reaches 80% or more. Studying the extensions added to each resource may provide guidance on missing data elements. (The data here also show that some of the existing native elements are unnecessary, or at least, utilized at very low levels, but removing existing data elements is more problematic than adding new ones.)
2. Focus initial anti-profiliferation efforts on the most frequently profiled resources, Extension and Observation. Also, examine methods for standardizing and reusing slices.
3. Create a curated registry for collecting, sorting, standardizing, and sharing reusable items. The registry should not only contain extensions, but also specialized identifiers, addresses, and other profiled datatypes that may be useful outside of the context of a full resource profile.
4. Within registries, provide better mechanisms for searching within StructureDefinitions to help users and tools find definitions with certain properties, such as the use of a certain code. Improve authoring tools to detect when authors have created a profile similar to an existing profile.
5. Since slicing contributes substantially to variation among profiles, it is worth exploring how slices could be shared and reused. For example, it might be possible to create additional complex datatypes derived from BackboneElement, like Timing or Dosage. This will allow standalone profiling of these elements.
6. Create a release cycle for the registry that leads to rapid publication of reusable items. The current practice of linking the official extension registry to the FHIR release cycle is far too slow for IG developers.
7. Follow up this study with a closer look at how Observations have been profiled and create better guidance on using this critical resource. Address the proper use Observation.component and point out alternative means for expressing ancillary information associated with observations. It may be helpful to create abstract profiles to serve as parent profiles for different types of observations. There could be more clarity around when to use a questionnaire, and when to use Observations with many components.
8. Strengthen education or governance that encourages IG creators to reuse existing extensions or create and contribute standalone extensions that have the best chance for reuse.
9. Educate IG developers on when to use fixed values, value patterns, and slicing approaches. Update old IGs and profiles to use value patterns when possible.
10. Conduct a similar study focused on the terminology landscape across a large sample of IGs.

## Conclusion

This paper was intended to shed light on how profiles are being used across the FHIR community by analyzing a diverse set of 125 implementation guides. Strong trends were revealed, including the concentration of profiling on a small number of resources and datatypes. Areas where current practice is creating non-reusable items and contributing to the proliferation of profiles were uncovered, and specific recommendations for creating more usable IGs were proposed.

The current study only examined StructureDefinitions, and a full study of profiling should include other conformance resources, especially those related to terminology. While the current study analyzed a large set of IGs, it might be possible to broaden the set, or focus on particular types of IGs, for additional insights.

## Data Availability

All data produced in the present study are available upon reasonable request to the authors

## Appendix A

List of Implementation Guides

IGs are listed by their formal identifiers. More information on each package can be found at https://registry.fhir.org/

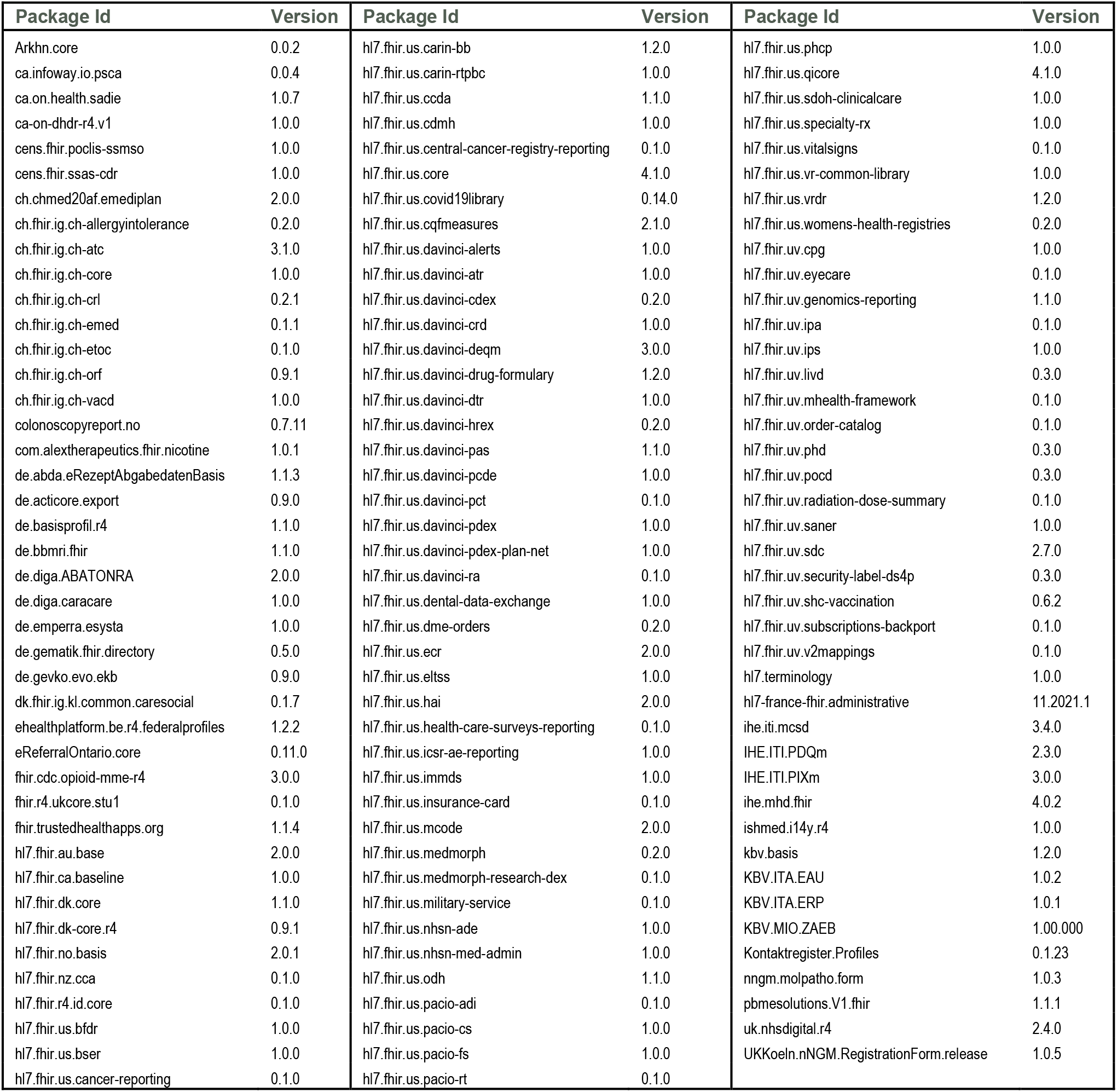

## Footnotes

1 To be fully transparent, FHIR 4.6.0 provides the extension elementdefinition-profile-element for the purpose of reusing a profiled BackboneElement. This is an obscure feature, not used in any IG analyzed here.

## References

.1 Health Level Seven. HL7 FHIR Release 4. [Online].; 2019 [cited 2022 February 22]. Available from: http://hl7.org/fhir/.

.1 Health Level Seven. [video].; 2019 [cited 2022 February 22]. Available from: https://www.youtube.com/watch?v=ovbRwbd0uD8.

.1 Health Level Seven. HL7 FHIR Release 4. [Online].; 2019 [cited 2022 February 22]. Available from: https://www.hl7.org/fhir/profiling.html.

.1 Firely. Simplifier.net. [Online]. [cited 2022 February 22]. Available from: https://simplifier.net/.

.1 Health Level Seven. FHIR Package Registry. [Online].; 2022 [cited 2022 February 22]. Available from: https://registry.fhir.org/.

.1 HL7 FHIR Foundation. FHIR Implementation Guide Registry. [Online].; 2017 [cited 2022 February 22]. Available from: http://fhir.org/guides/registry/.

.1 Health Level Seven. FHIR Resource StructureDefinition. [Online].; 2019 [cited 2022 February 22]. Available from: http://www.hl7.org/fhir/structuredefinition.html.

.1 Health Level Seven. FHIR Conformance Module. [Online].; 2019 [cited 2022 February 22]. Available from: https://www.hl7.org/fhir/conformance-module.html.

.1 Health Level Seven. FHIR Extensibility. [Online].; 2019 [cited 2022 February 22]. Available from: https://www.hl7.org/fhir/extensibility.html.

.1 Health Level Seven. FHIR Shorthand. [Online].; 2022 [cited 2022 February 22]. Available from: http://hl7.org/fhir/uv/shorthand/reference.html#contains-rules-for-extensions.

.1 Health Level Seven. HL7 FHIR Release 4. [Online].; 2019 [cited 2022 February 22]. Available from: https://www.hl7.org/fhir/patient.html.

.1 Health Level Seven. HL7 FHIR Release 4. [Online].; 2019 [cited 2022 February 22]. Available from: https://www.hl7.org/fhir/patient.profile.json.html.

.1 Health Level Seven. HL7 FHIR Release 5 Draft Ballot. [Online].; 2022 [cited 2022 February 22]. Available from: https://build.fhir.org/versions.html#maturity.

.1 Health Level Seven. HL7 FHIR Release 4. [Online].; 2019 [cited 2022 February 22]. Available from: https://hl7.org/fhir/2018May/conformance-rules.html#mustSupport.

.1 Greive G. Health Intersections. [Online].; 2014 [cited 2022 February 22]. Available from: http://www.healthintersections.com.au/?p=1924.

.1 Kavanagh R. FHIR Observations - design considerations, the proliferation of profiles, the role of validation [video].; 2018 [cited 2022 February 22]. Available from: https://www.youtube.com/watch?v=hLqjNfVyEa8.

.1 Health Level Seven. HL7 FHIR Release 4. [Online].; 2019 [cited 2022 February 22]. Available from: http://hl7.org/fhir/backboneelement.html#BackboneElement.

